# Rare *PANK2* Variants and Pantothenate Kinase-Associated Neurodegeneration in the Dominican Republic

**DOI:** 10.1101/2025.02.14.25321662

**Authors:** Badri N. Vardarajan, Pedro Sanchez-Rao, Christine Y. Kim, Peter Stoeter, Diones Rivera Mejia, Alexander Houck, Amanda Chan, Dolly Reyes-Dumeyer, Angel Piriz, Robert Fee, Francisco Blanco-Abinader, Elizabeth Rice, Samantha Christenson, Rebecca Chiu, Tamil I Gunasekaran, Rafael A. Lantigua, Clifton Dalgard, Serge Przedborski, Richard Mayeux

## Abstract

Pantothenate kinase-associated neurodegeneration (PKAN) is a rare, autosomal recessive neurological disorder characterized by the progressive degeneration of specific regions in the brain and is invariably fatal. Several individuals in families affected by PKAN were known to live in an isolated region in a southwestern province of the Dominican Republic and had been previously studied.

Forty-six individuals with PKAN in 34 families were evaluated for disease manifestations using the PKAN-Disease Rating Scale and the Leiter-3 Cognitive and Neuropsychological assessment. We completed whole genome sequencing in the 46 affected individuals and their 80 unaffected relatives. Haplotype analysis was used to identify shared genetic patterns among individuals with the mutation to identify common ancestral and founder effects.

The classical form of PKAN was observed in 22 individuals with moderate to severe oromandibular dystonia and limb dystonia and onset in early childhood. The atypical form was observed in 24 individuals with parkinsonism, dystonia, and cognitive impairment and later onset of disease. A *PANK2* variant, chr20:3907977: A:G *(c.680A>G, p.Y227C),* was homozygous among 42 affected individuals equally divided by disease form. There were 59 heterozygous carriers of this variant among parents and relatives of the affected individuals. Four individuals from two families were compound heterozygotes for *c.680A>G* and chr20:3918728: C:T (*c.1594C>T).* Haplotype analyses revealed shared patterns across families and of African origin consistent with founder effects for *c.680A>G* and *c.1594C>T*, likely introduced to the island 25 to 35 generations earlier. The frequency of heterozygous carriers of *c.680A>G* allele among individuals of Dominican ancestry living in New York was 0.18% but was 0.8% among individuals living in the Dominican Republic, significantly higher than the reported frequency for all causal *PANK2* mutations worldwide.

This investigation confirmed likely founder mutations in *PANK2* associated with the classical and atypical forms of PKAN in 34 families in an isolated region of the Dominican Republic. Compound heterozygosity was observed in four individuals from two families. The heterozygous frequency of *c.680A>G* was exceptionally high in the Dominican population compared with worldwide data. Founder mutations in such communities offer a unique opportunity to set up relevant, affordable and accessible genetic counseling and screening.

## Introduction

Pantothenate kinase-associated neurodegeneration (PKAN) is a rare, autosomal recessive neurological disorder characterized by the progressive degeneration in the globus pallidus and surrounding regions of the central nervous system.^1–3^ PKAN is the most common form of neurodegeneration associated with brain iron accumulation (NBIA), a group of clinical disorders characterized by abnormal involuntary movements, alterations in muscle tone, and extrapyramidal signs. Some, but not all these disorders, including PKAN, display characteristic radiographic evidence of iron accumulation in the brain, the “eye-of-the-tiger” sign, on magnetic resonance imaging.^3,4^

There are two forms of PKAN.^3^ Classical PKAN begins in early childhood and worsens gradually within the first five years of life. Dystonia of the musculature of the mouth, throat and tongue results in progressive dysarthria, dysphagia and eventually loss of speech and swallowing. There may be developmental delay and intellectual impairment, though the latter is difficult to assess due to limited verbal communication. In the classical form of PKAN, individuals experience a rapid course becoming wheelchair-bound by age 10 to 15 years, and the disease is invariably fatal.^5^ This form may also be associated with retinal degeneration and acanthocytosis.^6^ Atypical PKAN appears later in childhood or early adolescence, usually after the age of 10 years, and progresses more slowly than the classical form. In fact, the loss of ambulation may be delayed up as much as three decades.^5^ The characteristic manifestations include palilalia, dysarthria, parkinsonism, personality changes, and gait disorders. Impaired ambulation, mastication and dysphagia compound the overall survival with PKAN regardless of subtype. The phenotype of PKAN can also extend along a continuum without clear delineation between the classical and atypical forms, and there can be variable manifestations of age of onset and semiology.

PKAN is inherited as an autosomal recessive disease, caused by homozygous missense or loss of function mutations, or by compound heterozygous missense, completely penetrant, mutations in the *PANK2* gene.^1,3^ *PANK2* encodes the enzyme pantothenate kinase. Rare variants in this gene alter pantothenate, vitamin B5 metabolism, which is required to produce coenzyme A. Disruption of this enzyme affects energy and lipid metabolism leading to accumulation of potentially harmful compounds in the brain, including iron.^1^ *PANK2* is the only gene known to be associated with PKAN. The heterozygous frequency of *PANK2* variants has been estimated at less than 0.01% worldwide and varies slightly by ethnic group^7^, and the incidence of live births with PKAN is between 2 and 3 per 1,000,000 (0.0002%) worldwide. There are more than 100 pathogenic or likely pathogenic variants in *PANK2.*^8^

In 2022, we learned of several families affected by PKAN and living in an isolated region in the country, some of whom had been previously studied.^4,6^ Several years earlier 21 individuals had sequencing of the *PANK2* gene reporting a pathogenic variant, c.680A>G, in the homozygous configuration in 17 of these individuals.^4^ Four years later the variant was confirmed in this group of individuals and in another eight individuals.^6^ However, colleagues indicated that there might be several more affected individuals in this isolated region of approximately 30,000 individuals, and possibly more in other parts of the country. This meant that the heterozygous frequency of pathogenic *PANK2* alleles in this country might be higher than the worldwide frequency and raised the possibility of a founder mutation. They requested our assistance in setting up a genetic screening program.

## Materials and methods

In September 2023, we examined affected individuals with a clinical diagnosis of PKAN and their unaffected family members. We obtained blood for DNA and blood smear to assess acanthocytosis. Most individuals were evaluated at a hospital outpatient facility, but a small number that were unable to travel due to their disability were seen in their homes. A second set of families were evaluated in the capital in Santo Domingo. Most of the affected individuals had MRI brain imaging for diagnostic purposes.^4,6,9,10^

Our protocol was approved by the respective agencies in the Dominican Republic and the Institutional Review Board at Columbia University in accordance with the Declaration of Helsinki. All participating individuals or their designated authorized representative gave informed consent. Our assessments included a standardized medical history, medications used, a detailed family histories with pedigree construction, neuropsychological testing, and videotaping of a structured neurological examination that included the PKAN-Disease Rating Scale.^11^ With the help of the family, we attempted to determine the age at onset of symptoms by asking when they first noticed any sign or symptom possibly related to PKAN. We also asked when the affected individuals were last considered symptom free.

PKAN-Disease Rating Scale (PKAN-DRS).^11^ We assessed each individual in two phases. The first section rated cognition, behavior and disability using a modified Likert Scale from 0 (no impairment) to 4 (severe impairment). The behavioral assessment queried the presence of sadness, anxiety, suicidal thoughts, aggression, irritability, obsessions, and hallucinations. The disability assessment included speech, handwriting, feeding, eating, swallowing, hygiene, dressing, walking, schoolwork and vision. The second section rated motor manifestations using a similar Likert Scale approach and included: Parkinsonism (bradykinesia, rigidity, finger tapping, leg agility, freezing of gait, postural or truncal stability, and resting tremor), Dystonia (upper and lower face, eyes, jaw, tongue, neck, arms, trunk and legs), speech, gait, chorea, spasticity, action or postural tremor, and oculomotor dysfunction. The second section was videotaped for consensus review and scoring by a group of independent movement disorder specialists at Columbia University.

Leiter-3 Nonverbal Cognitive and Neuropsychological assessment.^12^ The Leiter-3 is a reliable and valid nonverbal test developed for individuals aged 3 to 75 years. The measure is specifically designed for use in individuals with cognitive, speech or hearing dysfunction, motor impairment, as well as individuals whose dominant language is not English. The measure uses a refined block-and-frame format with lightweight blocks and foam manipulatives to reduce the effects of impaired motor function, as well as visual stimuli (patterns, sequence, illustrations) that do not require the use of language.

Whole blood and DNA extraction. A blood sample was obtained for DNA extraction and collected from all individuals providing consent which included both affected and unaffected family members. Blood was shipped overnight to Columbia University in New York City where DNA was extracted and stored. An additional sample was sent to CEDIMAT in Santo Domingo to measure acanthocytosis by blood smear and examined under microscope to count the percentage of abnormally shaped cells.

Whole Genome Sequencing. To identify variants including single nucleotide variants (SNVs), indels, complex structural variation including insertions, deletions, copy number variation and tandem repeats, all affected individuals and their family members underwent whole genome sequencing PCR-free DNA library preparation was conducted by automation robotics ligation-based workflow. Quality controlled libraries were sequenced on the Illumina NovaSeq 6000 or X Plus at the Uniformed Services University of the Health Sciences in Bethesda, MD. Samples were sequenced to a depth of ≥30x mean coverage. Raw reads were aligned to the human reference genome build 38 using the Burrows–Wheeler Aligner.^13^ Quality control of the sequencing data and variant calling was done using Genome Analysis Toolkit (GATK).^14^ Called variants were further QCed using VariantRecalibration in GATK.

Sequence Analyses. Variants passing the Variant Quality Score Recalibration threshold were filtered for sample missingness (>2%), depth of coverage-DP>10 and genotype quality-GQ>20. We annotated high quality variants using ANNOVAR. Specifically, variants were annotated for population level frequency using Genome Aggregation Database (gnomAD)^15^, TOPMed and Database of Structural Variation (DGV)^16^, *in-silico* function using Ensembl Variant Effect Predictor (VEP)^17^ and variant conservation using Combined Annotation Dependent Depletion score (CADD)^18^ as well as variant-disease association from OMIM^19^, HGMD^20^ and ClinVar^21^. PKAN associated variants were expected to be significantly enriched in this cohort. We tested enrichment of putatively causal coding variants in the *PANK2* gene in the affected individuals in the cohort. Additionally, we tested the association of variants with disease severity and variants enriched among classical and atypical forms of PKAN.

Local ancestry was estimated using common variants defined with a minor allele frequency of less than 5% (MAF≥5%) in the 5 MB region flanking the *PANK2* gene. Haplotypes of affected and unaffected individuals were phased using Shapeit2 software.^22,23^ Reference haplotypes from three continental ancestries-non-Hispanic Whites (CEU), African (Yoruban) and native Americans (Pima, Sura, Maya and Incas) were obtained as previously described. Local ancestry proportions for each single nucleotide polymorphism (SNP) were determined using RFMix software^24^ and haplotype was assigned one of the three ancestries if at least 90% of the variants within the gene were of a specific ancestry.

## Results

We evaluated 126 individuals from 35 families, including 46 affected individuals equally distributed between the classical and atypical forms. The mean age of the affected individuals was 26.3 (± 10.4 s.d.) years, and the mean age at onset was 10.7 (± 3.8 s.d.) years (ranging from 2 to 20 years). However, symptom onset was uncertain for five individuals. For those classified as having the classical form, the mean age at onset was 8.1 (± 2.2 s.d.) years, while for those with the atypical form, it was 13.6 (± 2.7 s.d.) years. Girls or young women represented 51.1% of the affected individuals. On average, affected individuals had five years of education compared to nine years among the unaffected family members of similar age. Among the 75 family members, 49 (65%) were parents of the affected individuals, five were unaffected siblings, and the remaining 11 were unaffected more distant relatives.

Phenotypes. Among the 46 affected individuals, dystonia was the most prominent manifestation followed by parkinsonism (Table 1). Generalized dystonia was seen in most affected individuals. Facial dystonia was particularly prominent, either exclusively oromandibular or disproportionally oromandibular, with or without moderate blepharospasm. Mild to moderate bradykinesia was apparent in all affected individuals, but worse in those with prominent dystonia. The classical form of PKAN^3^, had onset between the ages of 3 and 10 years with rapid progression of symptoms, and was present in 54% of affected individuals. Clinically, these patients exhibited severe oromandibular dystonia, including buccolingual, dysarthria, and dysphagia. The atypical form of PKAN^3^, with onset after age 10 years in adolescence or adulthood, was present in 46% of affected individuals. This form was associated with a more gradual progression based on family report. We observed parkinsonism without tremor, namely bradykinesia and some rigidity, in most individuals with atypical PKAN. Two brothers carrying PKAN mutation, however, exhibited not only asymmetrical mild bradykinesia and rigidity, but also mild and intermittent resting tremor (i.e. tremor >1 cm but <3 cm in maximal amplitude) as well as shuffling gait, festination and freezing. Psychiatric symptoms, depression, anxiety, or behavioral abnormalities, and severe cognitive deficits were also prominent (Table 1). Visual deficits were suspected in a few subjects with either classical or atypical PKAN, but whether they had pigmentary retinopathy could not be readily established. In four affected individuals (8.7%), we could not determine the type of PKAN, but most had a later age at onset suggesting the atypical form. Acanthocytosis^6^ was present in 17 (39.5%) of the 43 affected individuals with PKAN who were tested.

Brain MRIs had been done previously in 41 of the affected individuals showing the classical findings of “eye-of-the-tiger” sign, characteristic of PKAN and other NBIA disorders (Table 1).^4,9,10^ The MRI protocol included diffusion-weighted and T1– and T2-weighted sequences. The findings included volume loss in the globus pallidus and midbrain nuclei^25,26^, and volume reduction in the medial frontal cortex^27^. In the white matter, the integrity of the tracts between the basal ganglia to the cerebellum and to the motor cortical areas was affected^26,28^ and resulted in a reduction in functional connectivity between these regions.^29^

Data acquired on the Leiter-3 scale determined the general cognitive functioning. Assessment of cognitive ability was not possible in 5 (11%) of the sample due to severe motor or cognitive limitations to perform and/or a poor understanding of instructions. The measure was completed in 41 individuals ranging in age from 7 to 54 and the overall nonverbal IQ for that group was in the very low/mildly delayed range (X̅=55.85, SD=16.62) with variability in individual subtest performance largely driven by reduced capacity.

Whole genome sequencing. A pathogenic variant, *PANK2* chr20:3907977:A:G *(c.680A>G, or p.Y227C* on Transcript: ENST00000316562.9) was present in the homozygous configuration among 42 affected individuals from 35 families. There were 59 heterozygous carriers of this variant, most of whom were parents or other relatives of the affected individuals. Interestingly, four affected individuals, of which three from one family were *PANK2* compound heterozygous: *c.680A>G* and chr20:3918728:C:T (*c.1594C>T or p.R422W on Transcript: ENST00000610179.7)*. This second missense variant has a Combined Annotation Dependent Depletion Score (CADD) of 26.4 that defines it as damaging^30^ and was extremely rare in gnomAD (MAF= 0.0000037) or other population databases. Three of the four were brothers from a single family (Figure 1). The other individual was an offspring of a *c.1594C>T* heterozygous carrier. An earlier report of this family was published as an abstract.^31^ All pathological variants were confirmed in a Clinical Laboratory Improvement Amendments (CLIA) laboratory enabling return results to physicians and their patients and families. We found no genetic differences to explain specific the disease phenotype, classical compared with atypical, nor age-at-onset.

**Figure 1.**
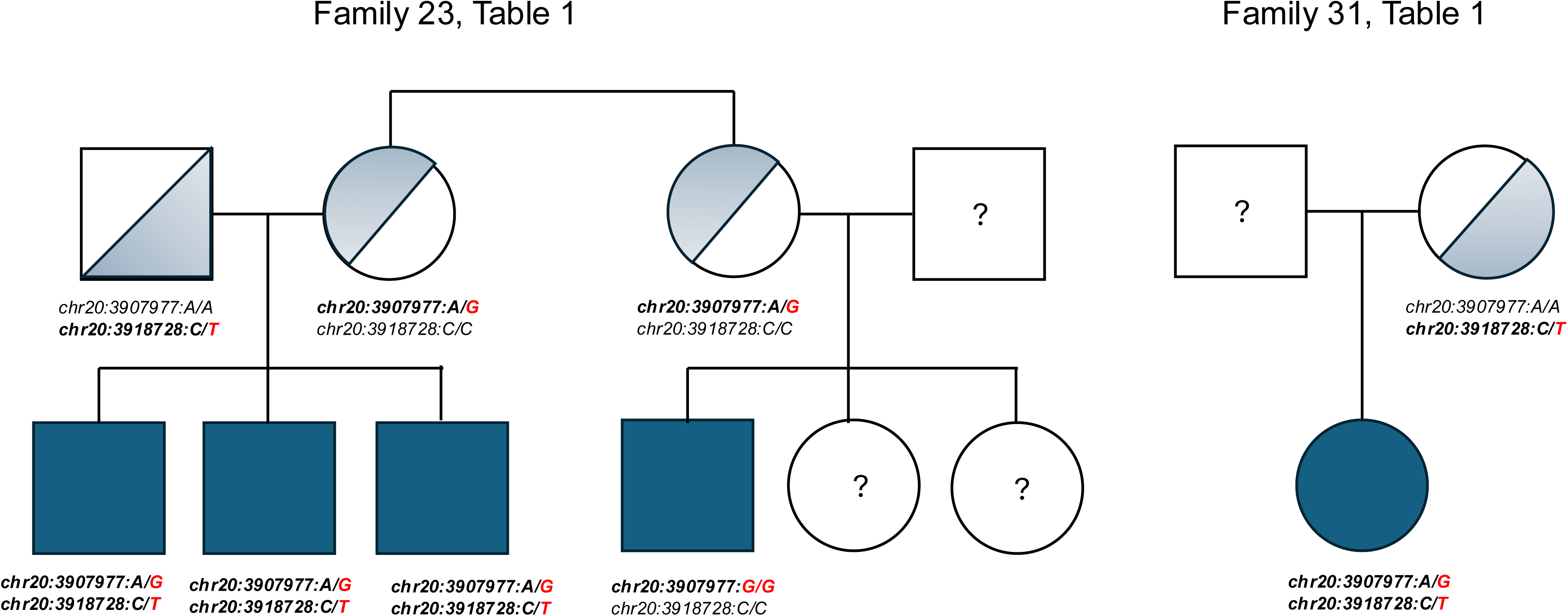
Legend. *PANK2* compound heterozygous variants in two families: chr20:3907977:A:G *(c.680A >G, p.Y227C)* and chr20:3918728:C:T (c.1594C>T, *p.R422W)*.

We used existing data from several sources to determine the heterozygous carrier frequency of the *c.680A>G*, *PANK2* variant. Whole exome sequencing data from the Institute of Genomic Medicine at Columbia University was reviewed in 7,127 individuals of Hispanic ancestry from the Dominican Republic and living in the communities of Washington Heights, Inwood and Hamilton Heights in New York City. None had PKAN and were participating in multiple genetic studies. We found 13 (0.18%) individuals were heterozygous carriers for *c.680A>G* allele. An additional 6,414 individuals of mixed Hispanic and African ancestry were also assessed. Ten (0.15%) were heterozygous carriers of the *c.680A>G* allele. We also had previously acquired whole genome sequencing data from 2,120 elderly individuals from 487 families without PKAN living in the Dominican Republic participating in a study of familial Alzheimer’s disease. Selecting one individual from each family, we identified four unrelated heterozygous carriers of the *PANK2 c.680A>G* allele yielding a carrier frequency of 0.82% which is approximately an 80-fold increase compared to the worldwide frequency of any pathologic or likely pathologic *PANK2* variants.^7^ At this observed frequency, we would expect the incidence rate of PKAN in the Dominican Republic to be one per 15,000 births, which is 100 times more prevalent than global estimates in African Americans and 25-32 times more prevalent than in non-Hispanic white and Hispanic populations.^7^ There were no heterozygous carriers of the *c.1594C>T* variant allele in PANK2 in any of these three cohorts, nor was it reported in the Genome Aggregation Database.^15^

The high frequency of the *PANK2 c.680A>G* allele in the Dominican Republic and among individuals of Dominican ancestry prompted us to investigate the ancestral origin of this mutation in this group of affected individuals and families. We used 15 common variants within the *PANK2* gene to determine the ancestry of the haplotype of mutation carriers and non-carriers. Among homozygous *PANK2 c.680A>G* allele affected individuals, 80.5% inherited haplotypes from an African ancestry on both chromosomes. The remaining affected individuals inherited at least one chromosome on an African background. Similarly, all compound heterozygous *PKAN* patients also had an African haplotype on both chromosomes. Remarkably, among relatives of affected individuals with *PKAN* who were heterozygous for *PANK2 c.680A>G* allele, 89.3% had at least one African ancestral haplotype within the *PANK2* gene.

Assuming that the *PANK2 c.680A>G* allele represents a founder mutation; we estimated the age of the variant allele using the haplotype described above in the 5 MB region flanking *PANK2* gene. We used the *Gamma method*^32^, which is based on ancestral segment lengths, to determine the genetic length of ancestral haplotypes shared between individuals carrying the mutation. The resulting estimate suggested that the homozygous founder *PANK2 c.680A>G* allele was likely introduced to the island 35.9 (95% CI 28.9 – 44.7) generations earlier.^32^

## Discussion

In an isolated region in the Dominican Republic, we identified 42 affected individuals homozygous for the chromosome 20 *c.680A>G* variant allele in *PANK2.* While *PANK2* had previously been sequenced in some individuals, this was the first whole genome assessment of these individuals and their family members.^4,6^ Four additional affected individuals, heterozygous for this variant allele and a second variant allele on chromosome 20 *c.1594C>T,* were also identified among a total of 35 families. Slightly more individuals had the classical form more than the atypical form in these families. The disease affected both sexes equally. Many affected individuals had previously been found to have the characteristic eye-of-the-tiger sign^4,9,10^ on brain MRI showing the well-known lesion in the globus pallidus, but not all had been sequenced. In addition, an involvement of the tracts between the basal ganglia and the cortical motor regions resulting in functional disconnection and hyperactivation of their motor areas, possibly due to a defect of this negative feedback loop was observed.^4,9,10^ Using genetic data from independent studies that included individuals of Hispanic ancestry from the Dominican Republic that did not have PKAN, we found the carrier frequency of the *c.680A>G* variant in *PANK2* to range for 0.18% to 0.82% which was significantly higher than all known mutations in *PANK 2* worldwide combined.^7^

Movement disorders are prominent features in PKAN, with dystonia and parkinsonism among the most frequently reported manifestations in both classical and atypical forms of PKAN.^2,3^ Our two identified *PANK2* mutations have essentially been found in patients from the Dominican Republic descent, and none of the 46 individuals form this cohort presented with a neurological phenotype that differed from that reported in patients carrying more common *PANK2* mutations.^2,3^ This is in keeping with genotype-phenotype correlation studies showing that among over 100 distinct *PANK2* mutations, none was associated with a unique neurological phenotype.^33,34^

Consistent with previous findings^2,3^, dystonia in classical PKAN was often multifocal or generalized. While limb dystonia was quite disabling, limiting the ability of individuals to perform daily tasks such as walking or using their hands, the most dramatic features were oromandibular dystonia characterized by involuntary tongue protrusion and jaw opening with severe drooling; one individual had prominent opisthotonus. In all affected individuals with oromandibular dystonia, it was never focal but rather associated with limb dystonia and additional cranial dystonia such as torticollis and, to a lesser extent, blepharospasm. In most, oromandibular dystonia was associated with severe dysphonia and dysarthria^35–37^, rendering speech unintelligible and impairing swallowing, which precluded feeding by mouth. Although severe cases of tongue protrusion dystonia may lead to self-injurious behavior, such as tongue and lip biting^37^, this was not observed in this cohort. Sensory tricks can be a hallmark of dystonia, but in individuals with PKAN this question has been infrequently examined. Previously reported sensory tricks in individuals with PKAN include touching the chin with one or both hands, humming, whistling or placing objects in mouth could mitigate the abnormal movements of the jaw and tongue.^38,39^ In this cohort, we did not formally test for sensory tricks; a review of the videos did not reveal any of the common sensory tricks, but in several patients using a small towel to absorb excess saliva, and in one placing a tooth brush in her mouth did lessen jaw opening and tongue protrusion.

In addition to dystonia, parkinsonism was present but generally milder than dystonia among individuals with classical PKAN. Mild bradykinesia was commonly observed in our classical PKAN cases, adding to the functional impairments caused by dystonia. Mild rigidity was present in some subjects. Among the other neurological features associated with PKAN^2,3^, we did not observe any classical cases with chorea or upper motor neuron signs. However, some subjects exhibited severe, painless muscle atrophy, particularly in the lower limbs, without evidence of fasciculations. This atrophy appeared to exceed what would be expected from disuse alone and contributed to marked weakness and restricted mobility. Additional diagnostic testing, including electromyography and nerve conduction velocity, was unavailable but could be considered in a next step study. A review of the literature on PKAN, only rarely mentions muscle atrophy as part of the clinical presentation in PKAN patients.^40^

Atypical presentations of PKAN add to its clinical complexity. For instance, in some cases, affected individuals, especially with late onset, exhibit parkinsonism as a predominant feature rather than dystonia.^2,3^ The systematic review from Botsford et al.^41^ analyzed 11 studies published between 1981 and 2018 that described PKAN patients presenting with parkinsonism, providing a valuable analysis about the clinical presentation of this association. All publications reported features consistent with atypical parkinsonism, including lack of asymmetry, coexisting dystonia, and poor levodopa responsiveness. However, one patient with atypical PKAN with parkinsonism was reported to show improvement in tremor following levodopa administration.^42^ Notably, two brothers with late-onset PKAN exemplify this atypical presentation, as both presented with marked parkinsonism, including freezing of gait, a symptom associated with Parkinson’s disease or atypical parkinsonism as well as PKAN; one individual had no associated dystonia at 23 years’ disease duration. These two individuals underscore the phenotypic heterogeneity of PKAN, with certain genetic or environmental modifiers potentially influencing the expression of either dystonia or parkinsonism. While both individuals were homozygous for the *c.680A>G* variant allele in *PANK2*, they were distinct from other individuals with atypical PKAN. A review of their sequencing data yielded no evidence of mutations in parkin or other known genes^43^, suggesting that this isolated parkinsonism, though unusual, is likely part of the PKAN neurological spectrum rather than a reflection of concomitant disorders.

Although not observed in the families reported here, Rohani et al.^44^ reported three individuals with atypical PKAN who presented with a tremor-dominant form of the disease, which followed a relatively benign course. Among these individuals, one had dystonic tremor and two had parkinsonian tremor. All three patients had homozygous mutations in the *PANK2* gene and displayed the characteristic “eye-of-the-tiger” sign on brain MRI. This underscores PKAN as a potential cause of tremor and emphasizes the need to consider the diagnosis of PKAN even in patients initially diagnosed with essential, dystonic, or parkinsonian tremor.

The cognitive phenotype in PKAN can be quite variable with performance ranging within the impaired range to within the normative average range. These results have greatly depended upon how conceptual abilities are measured given physical and potential language limitations. Deficits in specific cognitive domains including attention, executive functioning, and verbal and visual learning have also been reported in PKAN.^45^ Multifactorial contributions likely impacted cognitive performance within the current group of individuals examined here. Aside from genetic influence on brain development there are likely socioeconomic and physical limitations affected intellectual development. These contributors will need to be further examined to better understand genotype-phenotype correlations.

Several neurological disorders, including PKAN have been associated with acanthocytosis, and are also genetically defined diseases characterized by progressive degeneration of the basal ganglia and neuromuscular manifestations often accompanied by elevated levels of creatine kinase.^46,47^ Autosomal recessive chorea-acanthocytosis and X-linked McLeod syndrome all share psychiatric symptoms, cognitive impairment, peripheral neuropathy, myopathy and epilepsy. As is present in PKAN, affected individuals may also have facial and oromandibular dystonia, tics, parkinsonism and postural tremor. Chorea-acanthocytosis has been associated with loss of function variants in the *VPS13A* gene.^48^

Pantothenate kinase-associated neurodegeneration (PKAN, OMIM 234200), is the most common form of NBIA. It is an autosomal recessive resulting from rare pathogenic mutations in the *PANK2* gene (OMIM 606157).^3,34,49,50^ The Genome Aggregation Database has 66 variants listed as likely pathogenic or pathogenic^15^, but there are several hundred mutations described. Compound heterozygotes in *PANK2* are also frequently observed. It has been estimated that incidence rate of PKAN is two for every million live births^7^, but by using the heterozygous frequency we observed in this study the incidence rate would be much higher among individuals from the Dominican Republic. While estimates of the frequency are limited globally, PKAN is a rare Mendelian disorder. Clinical presentation includes dystonia, rigidity, bradykinesia, dysarthria, pigmentary retinopathy and dementia.^1–3^ Age of onset and progression for PKAN is variable: from early onset in the first decade of life with rapid progression to later onset with slower progression which may present as an adult-onset movement disorder.^2,51,52^

Though not fully understood, it has been suggested that variants in *PANK2* alter the function of mitochondrial pantothenate kinase 2 reducing coenzyme A (CoA) metabolism primarily in the basal ganglia, especially in the globus pallidus.^7,53^ Normally, PANK2 activates and maintains mitochondrial acyl carrier protein^2^. Once activated this protein provides support for fatty acid synthesis, iron-sulfur cluster biogenesis and electron transport. How a damaging variant in *PANK2* leads to the disease phenotype is not clear.

The cohort described here may be one the largest group of affected individuals with the same mutation in the homozygous configuration. The origin of the *c.680A>G* variant allele in *PANK2* is also of great interest. Haplotype analysis in which DNA sequence was examined around a specific variant of interest on chromosome 20, *wa*s used to identify shared genetic markers among affected individuals. This finding allowed us to infer a common ancestry or founder effect in order to track the inheritance of the mutation through generations.^54^ Individuals with a founder mutation will often share a large region of identical DNA around the mutated gene, the haplotype, can be traced back to the original founder. Such “founder mutations” are most readily identified in populations with a high degree of genetic homogeneity, like isolated communities^55^ such as those in the Barahona Province.

The genetic architecture of an isolated community can be influenced by the geographical region, population migration, selection pressure, and cultural practices, which influence the unique variants present in the specific population. The part of the country we visited is relatively isolated because it is surrounded by mountains and separated by a river. The application of genomics in community settings, such as this one in the Dominican Republic, provides an opportunity to identify founder variants associated with human disease. The identification and characterization of founder variants can have implications for understanding the causes of the disease, and in implementing social policies such as genetic testing enabling precise diagnosis and prevention in an attempt towards precision medicine.^56,57^

## Acknowledgements

The authors thank the medical staff at the Hospital Municipal de Cabral and the Los Centros de Diagnóstico y Medicina Avanzada y de Conferencias Médicas y Telemedicina (CEDIMAT). We also thank the individuals in the Barahona Province for organizing the transportation of patients and their families to the two clinics where these studies were conducted including: Celito Arias (Driver), Aracelis Mejia (Coordinator/volunteer), Ana Francisca Batista Cuevas (Nurse), Esperanza Cuevas Batista (Nurse), Damaris Feliz Cuevas (Doctor), Sol Juana Ines Feliz Urbaez (Nurse), Cesarina Torres (Neurologist) Lourdes Vargas (Phlebotomist). We acknowledge the work of Yihao Li at Columbia University in analysis of the genetic sequencing, and Francisco Roedan, Yaquiris de la Cruz, Joan Bonilla, also at Columbia University each of whom assisted in collecting clinical data and blood samples.

## Data Availability

Anonymous clinical and genetic data from this study are available by email request to the authors at Columbia University. The data are held on a secure website that also provides a Qualtrics form to be filled out by the investigators with contact information, title of the project, type of data requested, brief description of rational for the request, co-authors, research question to be addressed, and approval by institutional ethical review board.

---

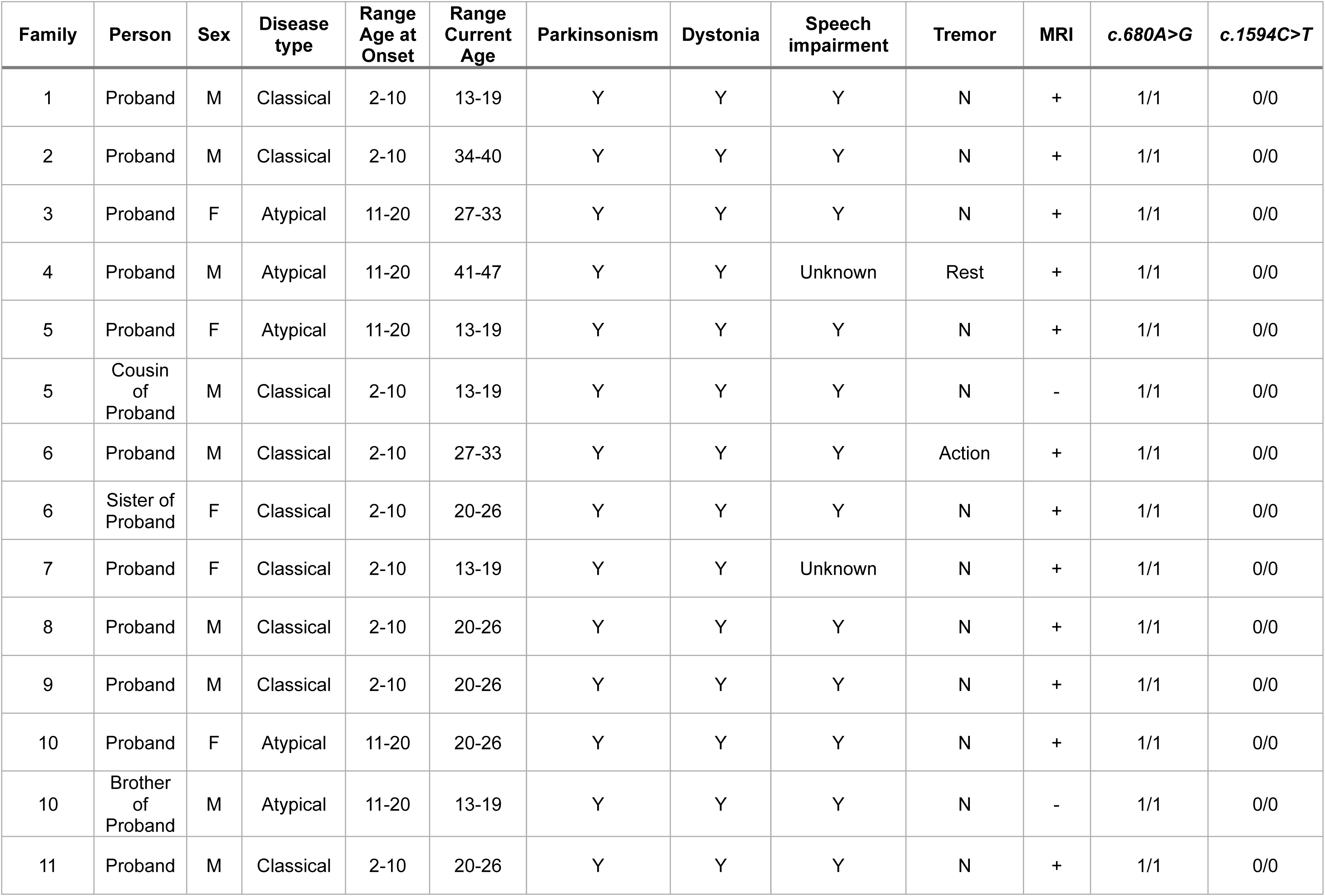
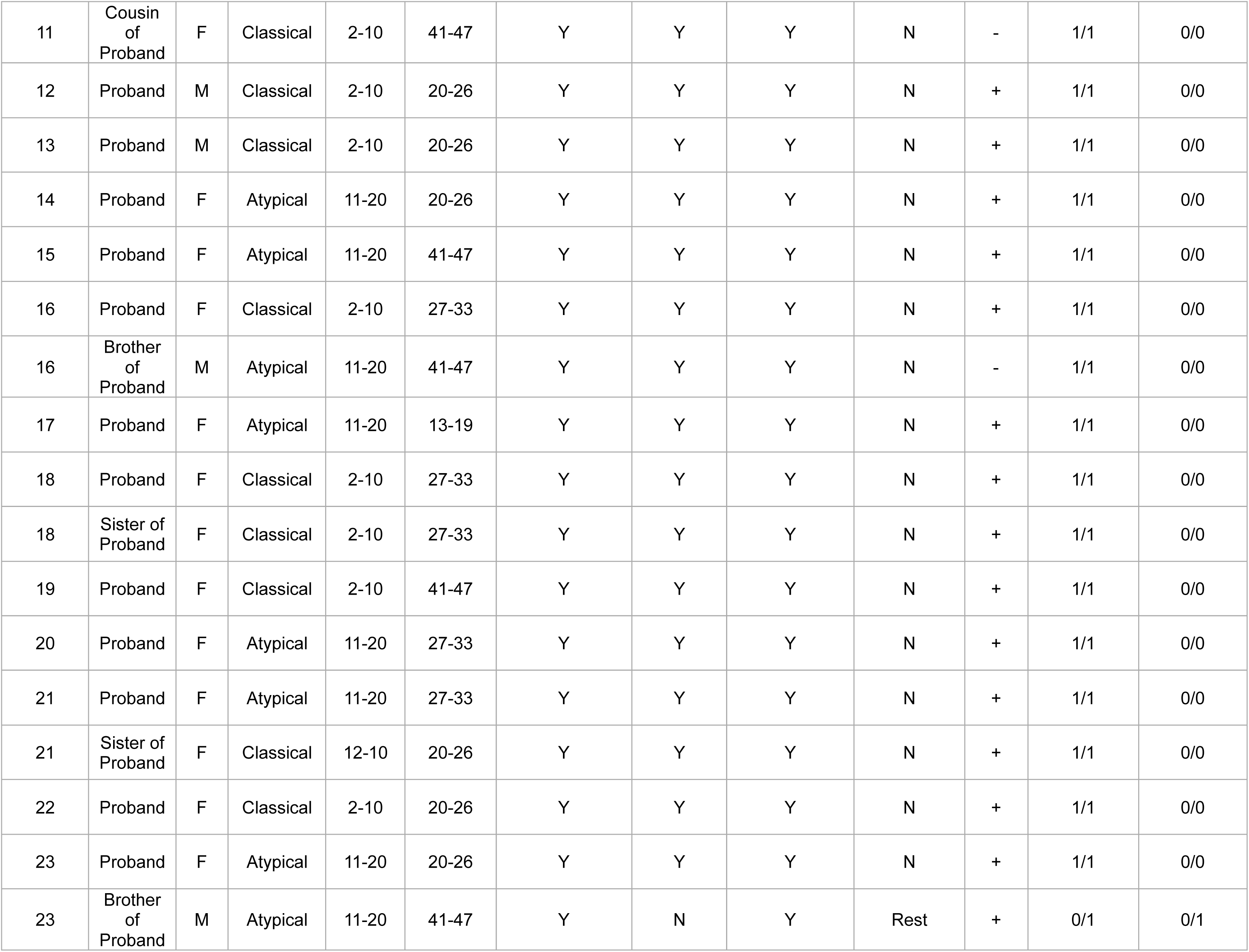
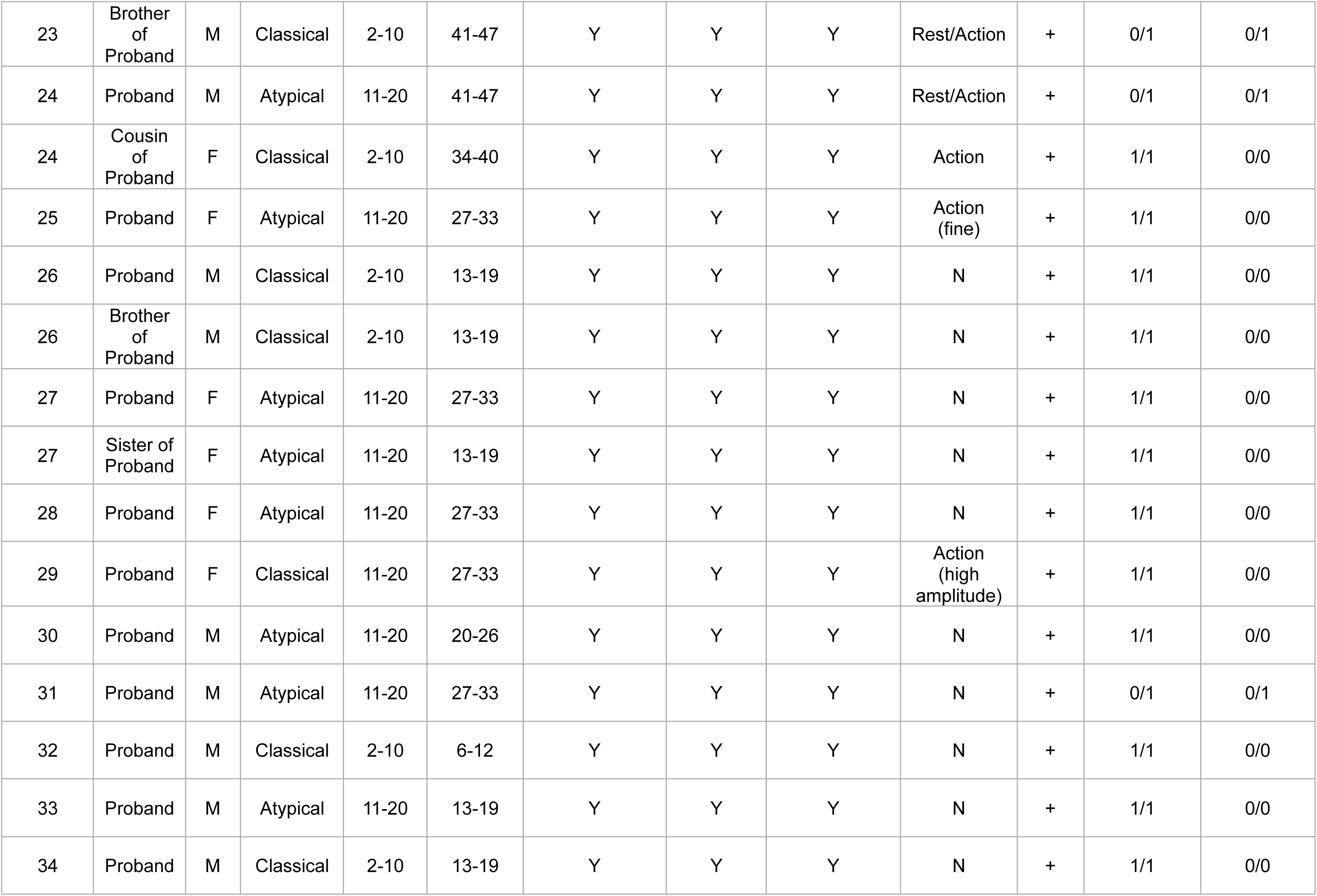
Phenotypic Description, PANK2 phenotypes of 46 Individuals with PKAN.

**Phenotypic Description, PANK2 phenotypes of 46 Individuals with PKAN.**
| Family | Person | Sex | Disease type | Range Age at Onset | Range Current Age | Parkinsonism | Dystonia | Speech impairment | Tremor | MRI | c.680A>G | c.1594C>T |
| --- | --- | --- | --- | --- | --- | --- | --- | --- | --- | --- | --- | --- |
| 1 | Proband | M | Classical | 2-10 | 13-19 | Y | Y | Y | N | + | 1/1 | 0/0 |
| 2 | Proband | M | Classical | 2-10 | 34-40 | Y | Y | Y | N | + | 1/1 | 0/0 |
| 3 | Proband | F | Atypical | 11-20 | 27-33 | Y | Y | Y | N | + | 1/1 | 0/0 |
| 4 | Proband | M | Atypical | 11-20 | 41-47 | Y | Y | Unknown | Rest | + | 1/1 | 0/0 |
| 5 | Proband | F | Atypical | 11-20 | 13-19 | Y | Y | Y | N | + | 1/1 | 0/0 |
| 5 | Cousin of Proband | M | Classical | 2-10 | 13-19 | Y | Y | Y | N | - | 1/1 | 0/0 |
| 6 | Proband | M | Classical | 2-10 | 27-33 | Y | Y | Y | Action | + | 1/1 | 0/0 |
| 6 | Sister of Proband | F | Classical | 2-10 | 20-26 | Y | Y | Y | N | + | 1/1 | 0/0 |
| 7 | Proband | F | Classical | 2-10 | 13-19 | Y | Y | Unknown | N | + | 1/1 | 0/0 |
| 8 | Proband | M | Classical | 2-10 | 20-26 | Y | Y | Y | N | + | 1/1 | 0/0 |
| 9 | Proband | M | Classical | 2-10 | 20-26 | Y | Y | Y | N | + | 1/1 | 0/0 |
| 10 | Proband | F | Atypical | 11-20 | 20-26 | Y | Y | Y | N | + | 1/1 | 0/0 |
| 10 | Brother of Proband | M | Atypical | 11-20 | 13-19 | Y | Y | Y | N | - | 1/1 | 0/0 |
| 11 | Proband | M | Classical | 2-10 | 20-26 | Y | Y | Y | N | + | 1/1 | 0/0 |
| 11 | Cousin of Proband | F | Classical | 2-10 | 41-47 | Y | Y | Y | N | - | 1/1 | 0/0 |
| 12 | Proband | M | Classical | 2-10 | 20-26 | Y | Y | Y | N | + | 1/1 | 0/0 |
| 13 | Proband | M | Classical | 2-10 | 20-26 | Y | Y | Y | N | + | 1/1 | 0/0 |
| 14 | Proband | F | Atypical | 11-20 | 20-26 | Y | Y | Y | N | + | 1/1 | 0/0 |
| 15 | Proband | F | Atypical | 11-20 | 41-47 | Y | Y | Y | N | + | 1/1 | 0/0 |
| 16 | Proband | F | Classical | 2-10 | 27-33 | Y | Y | Y | N | + | 1/1 | 0/0 |
| 16 | Brother of Proband | M | Atypical | 11-20 | 41-47 | Y | Y | Y | N | - | 1/1 | 0/0 |
| 17 | Proband | F | Atypical | 11-20 | 13-19 | Y | Y | Y | N | + | 1/1 | 0/0 |
| 18 | Proband | F | Classical | 2-10 | 27-33 | Y | Y | Y | N | + | 1/1 | 0/0 |
| 18 | Sister of Proband | F | Classical | 2-10 | 27-33 | Y | Y | Y | N | + | 1/1 | 0/0 |
| 19 | Proband | F | Classical | 2-10 | 41-47 | Y | Y | Y | N | + | 1/1 | 0/0 |
| 20 | Proband | F | Atypical | 11-20 | 27-33 | Y | Y | Y | N | + | 1/1 | 0/0 |
| 21 | Proband | F | Atypical | 11-20 | 27-33 | Y | Y | Y | N | + | 1/1 | 0/0 |
| 21 | Sister of Proband | F | Classical | 12-10 | 20-26 | Y | Y | Y | N | + | 1/1 | 0/0 |
| 22 | Proband | F | Classical | 2-10 | 20-26 | Y | Y | Y | N | + | 1/1 | 0/0 |
| 23 | Proband | F | Atypical | 11-20 | 20-26 | Y | Y | Y | N | + | 1/1 | 0/0 |
| 23 | Brother of Proband | M | Atypical | 11-20 | 41-47 | Y | N | Y | Rest | + | 0/1 | 0/1 |
| 23 | Brother of Proband | M | Classical | 2-10 | 41-47 | Y | Y | Y | Rest/Action | + | 0/1 | 0/1 |
| 24 | Proband | M | Atypical | 11-20 | 41-47 | Y | Y | Y | Rest/Action | + | 0/1 | 0/1 |
| 24 | Cousin of Proband | F | Classical | 2-10 | 34-40 | Y | Y | Y | Action | + | 1/1 | 0/0 |
| 25 | Proband | F | Atypical | 11-20 | 27-33 | Y | Y | Y | Action (fine) | + | 1/1 | 0/0 |
| 26 | Proband | M | Classical | 2-10 | 13-19 | Y | Y | Y | N | + | 1/1 | 0/0 |
| 26 | Brother of Proband | M | Classical | 2-10 | 13-19 | Y | Y | Y | N | + | 1/1 | 0/0 |
| 27 | Proband | F | Atypical | 11-20 | 27-33 | Y | Y | Y | N | + | 1/1 | 0/0 |
| 27 | Sister of Proband | F | Atypical | 11-20 | 13-19 | Y | Y | Y | N | + | 1/1 | 0/0 |
| 28 | Proband | F | Atypical | 11-20 | 27-33 | Y | Y | Y | N | + | 1/1 | 0/0 |
| 29 | Proband | F | Classical | 11-20 | 27-33 | Y | Y | Y | Action (high amplitude) | + | 1/1 | 0/0 |
| 30 | Proband | M | Atypical | 11-20 | 20-26 | Y | Y | Y | N | + | 1/1 | 0/0 |
| 31 | Proband | M | Atypical | 11-20 | 27-33 | Y | Y | Y | N | + | 0/1 | 0/1 |
| 32 | Proband | M | Classical | 2-10 | 6-12 | Y | Y | Y | N | + | 1/1 | 0/0 |
| 33 | Proband | M | Atypical | 11-20 | 13-19 | Y | Y | Y | N | + | 1/1 | 0/0 |
| 34 | Proband | M | Classical | 2-10 | 13-19 | Y | Y | Y | N | + | 1/1 | 0/0 |

## References

1. 1. Gregory A, Hayflick SJ. Pantothenate Kinase-Associated Neurodegeneration. In: Adam MP, Feldman J, Mirzaa GM, et al, eds. GeneReviews((R)). 1993.

2. Hayflick SJ, Jeong SY, Sibon OCM. PKAN pathogenesis and treatment. Mol Genet Metab. Nov 2022;137(3):283-291. doi:10.1016/j.ymgme.2022.09.011

3. Hayflick SJ, Westaway SK, Levinson B, et al. Genetic, clinical, and radiographic delineation of Hallervorden-Spatz syndrome. N Engl J Med. Jan 2 2003;348(1):33–40. doi:10.1056/NEJMoa020817

4. Delgado RF, Sanchez PR, Speckter H, et al. Missense PANK2 mutation without “eye of the tiger” sign: MR findings in a large group of patients with pantothenate kinase-associated neurodegeneration (PKAN). J Magn Reson Imaging. Apr 2012;35(4):788–94. doi:10.1002/jmri.22884

5. Amini E, Rohani M, Lang AE, et al. Estimation of Ambulation and Survival in Neurodegeneration with Brain Iron Accumulation Disorders. Mov Disord Clin Pract. Jan 2024;11(1):53–62. doi:10.1002/mdc3.13933

6. Schiessl-Weyer J, Roa P, Laccone F, et al. Acanthocytosis and the c.680 A>G Mutation in the PANK2 Gene: A Study Enrolling a Cohort of PKAN Patients from the Dominican Republic. PLoS One. 2015;10(4):e0125861. doi:10.1371/journal.pone.0125861

7. Brezavar D, Bonnen PE. Incidence of PKAN determined by bioinformatic and population-based analysis of ∼140,000 humans. Mol Genet Metab. Dec 2019;128(4):463–469. doi:10.1016/j.ymgme.2019.09.002

8. Landrum MJ, Lee JM, Benson M, et al. ClinVar: improving access to variant interpretations and supporting evidence. Nucleic Acids Res. Jan 4 2018;46(D1):D1062–D1067. doi:10.1093/nar/gkx1153

9. Fermin-Delgado R, Roa-Sanchez P, Speckter H, et al. Involvement of globus pallidus and midbrain nuclei in pantothenate kinase-associated neurodegeneration: measurement of T2 and T2* time. Clin Neuroradiol. Mar 2013;23(1):11–5. doi:10.1007/s00062-011-0127-9

10. Vilchez-Abreu C, Roa-Sanchez P, Fermin-Delgado R, et al. El signo del “Ojo del Tigre” en resonancia magnética: cambios relacionados con la edad. Anal Radiol Mexico. 2013;3:189–196.

11. Darling A, Tello C, Marti MJ, et al. Clinical rating scale for pantothenate kinase-associated neurodegeneration: A pilot study. Mov Disord. Nov 2017;32(11):1620–1630. doi:10.1002/mds.27129

12. Roid GH, Koch C. Leiter-3: Nonverbal Cognitive and Neuropsychological Assessment. In: McCallum R, ed. Handbook of Nonverbal Assessment. Springer, Cham; 2017.

13. Li H, Durbin R. Fast and accurate short read alignment with Burrows-Wheeler transform. Bioinformatics. Jul 15 2009;25(14):1754–60. doi:10.1093/bioinformatics/btp324

14. McKenna A, Hanna M, Banks E, et al. The Genome Analysis Toolkit: a MapReduce framework for analyzing next-generation DNA sequencing data. Genome research. Sep 2010;20(9):1297–303. doi:10.1101/gr.107524.110

15. Gudmundsson S, Singer-Berk M, Watts NA, et al. Variant interpretation using population databases: Lessons from gnomAD. Hum Mutat. Aug 2022;43(8):1012–1030. doi:10.1002/humu.24309

16. MacDonald JR, Ziman R, Yuen RK, Feuk L, Scherer SW. The Database of Genomic Variants: a curated collection of structural variation in the human genome. Nucleic Acids Res. Jan 2014;42(Database issue):D986–92. doi:10.1093/nar/gkt958

17. McLaren W, Gil L, Hunt SE, et al. The Ensembl Variant Effect Predictor. Genome Biol. Jun 6 2016;17(1):122. doi:10.1186/s13059-016-0974-4

18. Kircher M, Witten DM, Jain P, O’Roak BJ, Cooper GM, Shendure J. A general framework for estimating the relative pathogenicity of human genetic variants. Nat Genet. Mar 2014;46(3):310–5. doi:10.1038/ng.2892

19. McKusick-Nathans_Institute_of_Genetic_Medicine. Online Mendelian Inheritance in Man. Johns Hopkins University. http://omim.org/

20. Stenson PD, Ball EV, Mort M, et al. Human Gene Mutation Database (HGMD): 2003 update. Hum Mutat. Jun 2003;21(6):577–81. doi:10.1002/humu.10212

21. ClinVar. ClinVar Database. www.ncbi.nlm.nih.gov/clinvar

22. Delaneau O, Marchini J, Zagury JF. A linear complexity phasing method for thousands of genomes. Nat Methods. Dec 4 2011;9(2):179–81. doi:10.1038/nmeth.1785

23. Koenig Z, Yohannes MT, Nkambule LL, et al. A harmonized public resource of deeply sequenced diverse human genomes. bioRxiv. Feb 28 2024; doi:10.1101/2023.01.23.525248

24. Maples BK, Gravel S, Kenny EE, Bustamante CD. RFMix: a discriminative modeling approach for rapid and robust local-ancestry inference. Am J Hum Genet. Aug 8 2013;93(2):278–88. doi:10.1016/j.ajhg.2013.06.020

25. Roa-Sanchez P, Bido P, Oviedo J, Huppertz HJ, Speckter H, Stoeter P. Changes in Cerebral Gray and White Matter in Patients with Pantothenate Kinase-Associated Neurodegeneration: A Long-Term Magnetic Resonance Imaging Follow-Up Study. J Mov Disord. May 2021;14(2):148–152. doi:10.14802/jmd.20102

26. Stoeter P, Roa P, Foerster B, et al. Volumes of midbrain nuclei in Pantothenate-Kinase Associated Neurodegeneration (PKAN) Dystonia. ARC J Radiol Med Imaging 2020;4:19–25.

27. Rodriguez-Raecke R, Roa-Sanchez P, Speckter H, et al. Grey matter alterations in patients with Pantothenate Kinase-Associated Neurodegeneration (PKAN). Parkinsonism Relat Disord. Sep 2014;20(9):975–9. doi:10.1016/j.parkreldis.2014.06.005

28. Rivera D, Roa-Sanchez P, Bido P, Speckter H, Oviedo J, Stoeter P. Cerebral and cerebellar white matter tract alterations in patients with Pantothenate Kinase-Associated Neurodegeneration (PKAN). Parkinsonism Relat Disord. May 2022;98:1–6. doi:10.1016/j.parkreldis.2022.03.017

29. Stoeter P, Roa P, Bido P, Speckter H, Oviedo J, Rodriguez-Raecke R. Functional connectivity of the motor system in dystonia due to PKAN. eNeurologicalSci. Mar 2021;22:100314. doi:10.1016/j.ensci.2021.100314

30. Schubach M, Maass T, Nazaretyan L, Roner S, Kircher M. CADD v1.7: using protein language models, regulatory CNNs and other nucleotide-level scores to improve genome-wide variant predictions. Nucleic Acids Res. Jan 5 2024;52(D1):D1143–D1154. doi:10.1093/nar/gkad989

31. Middleton F, Muniz C, Ojukwu I, et al. Phenotypic analysis of subjects with homozygous and compound heterozygous PANK2 mutations in a single extended pedigree from the Dominican Republic (DR) PKAN cohort. Mov Disord. 2020;35(Supplement 1)

32. Gandolfo LC, Bahlo M, Speed TP. Dating rare mutations from small samples with dense marker data. Genetics. Aug 2014;197(4):1315–27. doi:10.1534/genetics.114.164616

33. Chang X, Zhang J, Jiang Y, Wang J, Wu Y. Natural history and genotype-phenotype correlation of pantothenate kinase-associated neurodegeneration. CNS Neurosci Ther. Jul 2020;26(7):754–761. doi:10.1111/cns.13294

34. Hartig MB, Hortnagel K, Garavaglia B, et al. Genotypic and phenotypic spectrum of PANK2 mutations in patients with neurodegeneration with brain iron accumulation. Ann Neurol. Feb 2006;59(2):248–56. doi:10.1002/ana.20771

35. Esper CD, Freeman A, Factor SA. Lingual protrusion dystonia: frequency, etiology and botulinum toxin therapy. Parkinsonism Relat Disord. Aug 2010;16(7):438–41. doi:10.1016/j.parkreldis.2010.04.007

36. Saeedi Y, Kazemi F, Habibi SAH, et al. Tongue Protrusion Dystonia in Pantothenate Kinase-Associated Neurodegeneration. Pediatr Neurol. Feb 2020;103:76–78. doi:10.1016/j.pediatrneurol.2019.06.004

37. Schneider SA, Aggarwal A, Bhatt M, et al. Severe tongue protrusion dystonia: clinical syndromes and possible treatment. Neurology. Sep 26 2006;67(6):940–3. doi:10.1212/01.wnl.0000237446.06971.72

38. Martins J, Darling A, Garrido C, et al. Sensory Tricks in Pantothenate Kinase-Associated Neurodegeneration: Video-Analysis of 43 Patients. Mov Disord Clin Pract. Nov 2019;6(8):704–707. doi:10.1002/mdc3.12842

39. Petrovic IN, Kresojevic N, Ganos C, et al. Characteristic “Forcible” Geste Antagoniste in Oromandibular Dystonia Resulting From Pantothenate Kinase-Associated Neurodegeneration. Mov Disord Clin Pract. Jun 2014;1(2):112–114. doi:10.1002/mdc3.12035

40. Pan S, Zhu C. Atypical pantothenate kinase-associated neurodegeneration with PANK2 mutations: clinical description and a review of the literature. Neurocase. Jun 2020;26(3):175–182. doi:10.1080/13554794.2020.1752739

41. Botsford E, George J, Buckley EE. Parkinson’s Disease and Metal Storage Disorders: A Systematic Review. Brain Sci. Oct 31 2018;8(11)doi:10.3390/brainsci8110194

42. Feuerstein J, Olvera C, Fullard M. Treatment Responsiveness of Parkinsonism in Atypical Pantothenate Kinase-Associated Neurodegeneration. Mov Disord Clin Pract. Sep 2020;7(Suppl 3):S71–S73. doi:10.1002/mdc3.13056

43. Riboldi GM, Frattini E, Monfrini E, Frucht SJ, Di Fonzo A. A Practical Approach to Early-Onset Parkinsonism. J Parkinsons Dis. 2022;12(1):1–26. doi:10.3233/JPD-212815

44. Rohani M, Shahidi G, Alavi A, et al. Tremor-Dominant Pantothenate Kinase-associated Neurodegeneration. Mov Disord Clin Pract. Sep-Oct 2017;4(5):772–774. doi:10.1002/mdc3.12512

45. Freeman K, Gregory A, Turner A, Blasco P, Hogarth P, Hayflick S. Intellectual and adaptive behaviour functioning in pantothenate kinase-associated neurodegeneration. J Intellect Disabil Res. Jun 2007;51(Pt. 6):417–26. doi:10.1111/j.1365-2788.2006.00889.x

46. Peikert K, Danek A, Hermann A. Current state of knowledge in Chorea-Acanthocytosis as core Neuroacanthocytosis syndrome. Eur J Med Genet. Nov 2018;61(11):699–705. doi:10.1016/j.ejmg.2017.12.007

47. Vaisfeld A, Bruno G, Petracca M, et al. Neuroacanthocytosis Syndromes in an Italian Cohort: Clinical Spectrum, High Genetic Variability and Muscle Involvement. Genes (Basel). Feb 26 2021;12(3)doi:10.3390/genes12030344

48. Chaudhari S, Ware AP, Jasti DB, et al. Exome sequencing of choreoacanthocytosis reveals novel mutations in VPS13A and co-mutation in modifier gene(s). Mol Genet Genomics. Jul 2023;298(4):965–976. doi:10.1007/s00438-023-02032-2

49. Pellecchia MT, Valente EM, Cif L, et al. The diverse phenotype and genotype of pantothenate kinase-associated neurodegeneration. Neurology. May 24 2005;64(10):1810–2. doi:10.1212/01.WNL.0000161843.52641.EC

50. Zhou B, Westaway SK, Levinson B, Johnson MA, Gitschier J, Hayflick SJ. A novel pantothenate kinase gene (PANK2) is defective in Hallervorden-Spatz syndrome. Nat Genet. Aug 2001;28(4):345–9. doi:10.1038/ng572

51. Ma LY, Wang L, Yang YM, Lu Y, Cheng FB, Wan XH. Novel gene mutations and clinical features in patients with pantothenate kinase-associated neurodegeneration. Clin Genet. 2015;87(1):93–5. doi:10.1111/cge.12341

52. Thomas M, Hayflick SJ, Jankovic J. Clinical heterogeneity of neurodegeneration with brain iron accumulation (Hallervorden-Spatz syndrome) and pantothenate kinase-associated neurodegeneration. Mov Disord. Jan 2004;19(1):36–42. doi:10.1002/mds.10650

53. Jeong SY, Hogarth P, Placzek A, et al. 4’-Phosphopantetheine corrects CoA, iron, and dopamine metabolic defects in mammalian models of PKAN. EMBO Mol Med. Dec 2019;11(12):e10489. doi:10.15252/emmm.201910489

54. Andres AM, Nowick K. Editorial overview: genetics of human evolution: the genetics of human origins. Curr Opin Genet Dev. Dec 2014;29:v–vii. doi:10.1016/j.gde.2014.11.001

55. Peltonen L, Palotie A, Lange K. Use of population isolates for mapping complex traits. Nat Rev Genet. Dec 2000;1(3):182–90. doi:10.1038/35042049

56. Sandler S, Alfino L, Saleem M. The importance of preventative medicine in conjunction with modern day genetic studies. Genes Dis. Jun 2018;5(2):107–111. doi:10.1016/j.gendis.2018.04.002

57. Zlotogora J, Carmi R, Lev B, Shalev SA. A targeted population carrier screening program for severe and frequent genetic diseases in Israel. Eur J Hum Genet. May 2009;17(5):591–7. doi:10.1038/ejhg.2008.241

