## Supplementary figures and images for "Rare *PANK2* Variants and Pantothenate Kinase-Associated Neurodegeneration in the Dominican Republic"

### Supplemental Figure 1. Cabral. Dom Repub.pptx

#### Slide 1
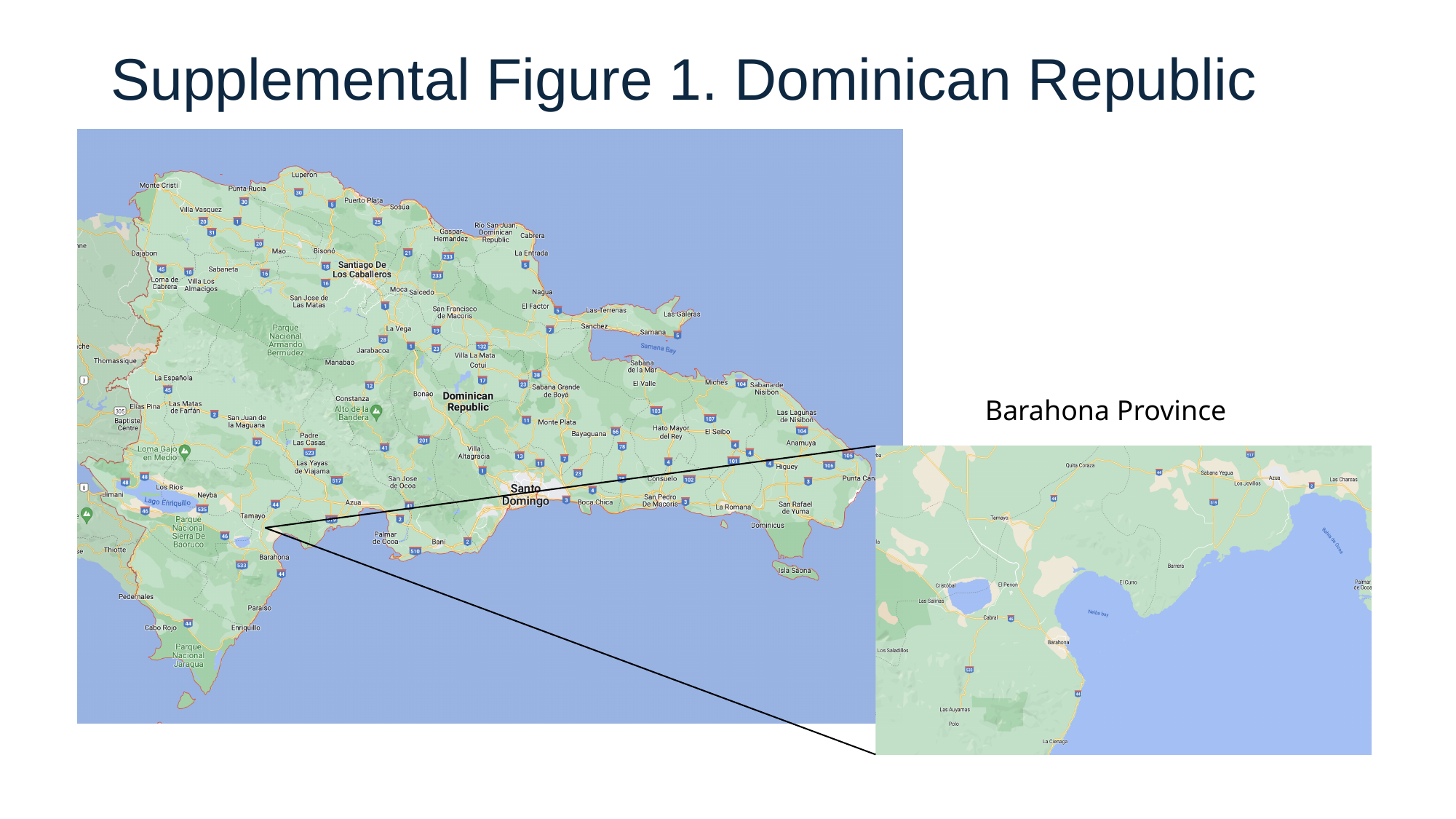

##### Supplemental Figure 1. Dominican Republic
Barahona Province
